# Verbal Autopsy Manager (VMan3): A comprehensive software tool for managing and improving quality, availability and use of cause of death data from community deaths

**DOI:** 10.1101/2025.05.28.25328220

**Authors:** Isaac Lyatuu, Collins Odhiambo, Sigilbert Mrema, Jonas Mwambimbi, Chriss Kimaryo, Shoko Irema, Saviour Moyo, Gisbert Msigwa, Godfrey Kayita, Robert Mswia

## Abstract

Accurate mortality and cause-of-death (COD) data are critical for informing health policy, guiding resource allocation, and supporting epidemiological surveillance. However, many low- and middle-income countries (LMICs) face persistent challenges in reporting deaths and determining causes of death for deaths occurring outside health facilities, largely due to the absence of robust community-level reporting systems. Verbal Autopsy (VA), which involves interviewing the deceased’s relatives or caregivers to infer COD, provides a feasible alternative in such contexts. Ensuring high-quality data collection and management is essential to derive valid COD outcomes from VAs. This study presents the development of Verbal Autopsy Manager Version 3 (VMan3) - a comprehensive software platform designed to improve the management, analysis, and quality of VA data. VMan3 supports digital COD assignment, integrated data quality assessment, and interoperability with national health information systems. The platform features a modular architecture that supports future scalability. Its backend is developed using Python’s FastAPI framework for high-performance API integration, while the frontend is built with AngularJS to deliver a responsive and interactive user experience. Data storage and retrieval are handled through the ArangoDB open-source database. The initial release of VMan3 includes features such as a dashboard for visualizing VA submissions by month, sex, and age group (neonates, children, adults), a geospatial module for mapping interview locations, a computer-coded verbal autopsy (CCVA) module for automated COD assignment, a physician-coded verbal autopsy (PCVA) module supporting online physician review, a data quality module to assess and improve reliability, and a system settings module for customization and configuration. VMan3 represents a significant step toward strengthening mortality surveillance and data-driven decision-making in LMICs

**Author Summary:** VA instrument consists of structured and semi-structured questions designed to collect information on the deceased’s signs and symptoms, medical history, demographic characteristics, and care-seeking behavior prior to death. Depending on the specific circumstances surrounding each death, the length and complexity of the VA interview can vary, resulting in records with diverse depth and structure. This variability presents challenges for data management, consistency, and quality. Verbal Autopsy Manager Version 3 (VMan3) addresses these challenges by utilizing a document-oriented data model, which allows flexible, schema-less handling of VA records. This approach accommodates the complex and variable nature of VA data, improving the user experience and enabling efficient processing and analysis. VMan3 supports both automated (computer-coded verbal autopsy, CCVA) and physician-led (physician-coded verbal autopsy, PCVA) cause-of-death assignment, while its configurable system settings allow easy adaptation to country-specific needs and workflows. We recommend the adoption of VMan3 by countries and programs implementing VA to strengthen data collection processes and enhance the completeness, accuracy, and availability of mortality statistics essential for informed public health planning and policy.

## Introduction

Accurate death and cause of death (COD) information is essential for informing policy and decision-making, resource allocation, health system performance assessment, mortality trend analysis, public health emergency (PHE) detection and supporting targeted health interventions. Countries must have the capacity to routinely generate and utilize high-quality mortality and COD data to support these functions. However, many countries in the World Health Organization’s African Region (WHO-AFRO) struggle with systematically producing and using reliable mortality data (1,2). These limitations were particularly evident during the COVID-19 pandemic when countries in the WHO-AFRO region reported only 155,248 of an estimated 439,500 COVID-19 deaths (3), highlighting gaps in real-time mortality surveillance and reporting systems.

Recognizing these challenges, the “Decade on Civil Registration” (2015–2024) in Africa spurred numerous Civil Registration and Vital Statistics (CRVS) improvement initiatives. Many countries have updated legal frameworks, adopted international standards, trained personnel, launched e-learning and capacity-building programs, and, for the first time, published vital statistics reports. These efforts present a unique opportunity to collaborate with academic and research institutions to provide technical expertise in strengthening and maintaining CRVS systems. Key areas of potential support include system assessments, data analysis, monitoring and evaluation, capacity development, and implementation research.

Despite these advancements, over two-thirds of deaths worldwide still lack a documented cause, with the majority occurring in low-and middle-income countries (LMICs). Limited resources, weak health infrastructure, and insufficient expertise hinder comprehensive COD reporting. To bridge this gap, **Verbal Autopsy (VA)** has emerged as an effective method for determining COD for community deaths by systematically interviewing family members or caregivers of the deceased (4). VA collects information on symptoms and circumstances leading to death, which is then analyzed by physicians or automated computer algorithms to infer the most likely cause.

VA interviews are typically conducted by non-medical personnel using standardized, semi-structured questionnaires. The WHO has developed guidelines and tools (5) to facilitate VA data collection and analysis, ensuring consistency across countries. With increasing efforts to improve CRVS systems, several WHO-AFRO nations have adopted VA to enhance the availability of mortality data (4). Many of these countries have strengthened their legal and institutional frameworks, established teams for vital event reporting, and integrated VA into their mortality surveillance programs. This has expanded VA’s role in public health decision-making, increasing the demand for high-quality VA data to inform policy, planning and PHE management.

The WHO, through the Verbal Autopsy Reference Group (VARG), supports the implementation of VA by developing standardized questionnaires, refining COD algorithms, training users, and providing guidelines for data management, analysis, and dissemination. These collective efforts aim to improve VA data quality and promote its integration into national mortality reporting frameworks.

The growing availability of VA data highlights the need for rigorous data quality control and effective utilization. Ensuring high-quality VA data requires a meticulous focus on completeness, consistency, and adherence to standardized data collection protocols. Moreover, linking VA data with health and civil registration datasets is critical to achieving comprehensive mortality coverage. Standardized data collection and integration facilitate cross-country comparisons, enabling policymakers to derive meaningful insights and address broader regional and global health challenges. Strengthening VA systems and data management practices will ultimately enhance mortality surveillance and improve health outcomes in LMICs.

### Verbal Autopsy Software Tools

The implementation of VA has been further supported by the development of dedicated software tools, such as the Verbal Autopsy Manager (VMan) (6), OpenVA (7), SmartVA (8) and the Verbal Autopsy Tool (VATool) (9). Developed in Tanzania and Kenya, respectively, these tools have been successfully implemented in Tanzania, Kenya, Zambia, Ethiopia, and Rwanda, improving VA data collection, processing, and reporting. Other VA software solutions, including VA-Explorer (10) and easyVA (developed with support from the Bill & Melinda Gates Foundation), also offer similar functionalities.

These VA tools enable structured data management, processing, visualization, and integration into national health systems. VMan, for instance, provides features such as online physician coding using International Classification of Diseases (ICD) standards, automated COD classification using existing VA algorithms, and dynamic data visualization. However, challenges remain regarding configuration flexibility, user authentication mechanisms, system interoperability, automation of COD assignment and processing of large volumes of VA data. Addressing these challenges is crucial for optimizing VA tools and ensuring their seamless integration into broader mortality surveillance frameworks.

This project presents the development of the Verbal Autopsy Manager Version 3 (VMan3), a software tool designed to address some challenges with existing software, enhance VA data quality, automate COD classification, and facilitate integration with national and global health information systems.

## Methods

### System Design

The VMan3 software tool is structured around a three-tier architecture, emphasizing configurability, scalability, and interoperability. The development followed an agile methodology, ensuring continuous user engagement, validation, and adaptability. In phase 1 of planning, multiple stakeholders from Zambia and Tanzania were engaged to capture key needs and features expected for VA software. This was done interactively in Zambia where stakeholders’ needs were elicited through sticky notes, active mapping of sticky notes ideas to system modules, prioritization of system features and discussions. In phase 2 of system design, the development team considered feedback from the output of phase 1 to design a system architecture that captures the stakeholder needs with prioritized system features as shown in figure 1. Phase 3 system development and phase 4 system testing occurred iteratively with the development team and testing team working together to ensure each module added during development is tested. The system was deployed in the cloud with dummy data to get more user testing and feedback from stakeholders and in-country VA implementation teams. The dummy data was downloaded from a test instance of Open Data Kit (ODK) Central with over 40,000 VA records. The records were processed and visualized in the dashboard.

**Figure 1:**
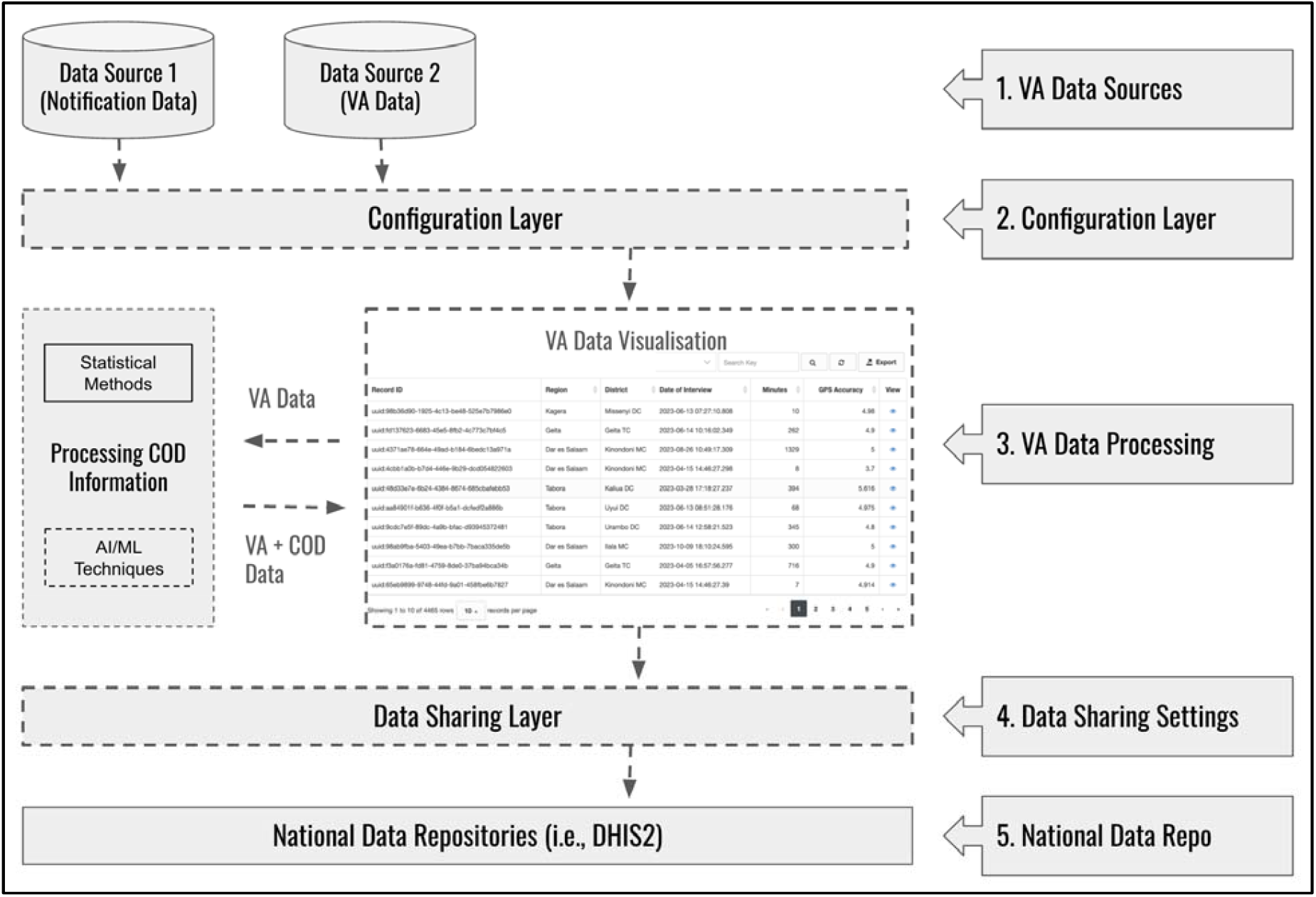
Verbal Autopsy Manager design architecture

### Configurable Design

Allows customization to accommodate country-specific administrative structures and integrates with various VA data sources, such as ODK Collect and Central. This flexibility ensures adherence to standardized VA protocols while allowing tailored implementations.

### Scalable Infrastructure

An architecture that supports expansion across multiple regions and levels of health system implementation, ensuring efficient data processing even as mortality surveillance scales up.

### Interoperability for Data Integration

The use of standardized Application Programming Interfaces (APIs) to facilitate seamless data exchange with health information and civil registration systems, ensuring consistency and accessibility of VA and cause of death data.

## Results

### Dashboard Module (Figure 2)

The dashboard module contains standard graphs and tables that present overview of the current collected data. This includes information on total submission by month and year, gender and the predefined age groups in the WHO-VA questionnaire. In addition, if the Cause of Death (COD) is available (COD requires prior processing of either CCVA or PCVA), the distribution of cause specific mortality fractions by age group (neonate, children and adults) and male and female is presented on the second row of the dashboard. The third row contains aggregate information of the distribution of VA data b the administrative levels as defined in the settings page. The dashboard module also contains a global filter functionality on the top, which allows the view to filter VA records based on date (reporting date, submission date or date of death) and location of the interview.

**Figure 2:**
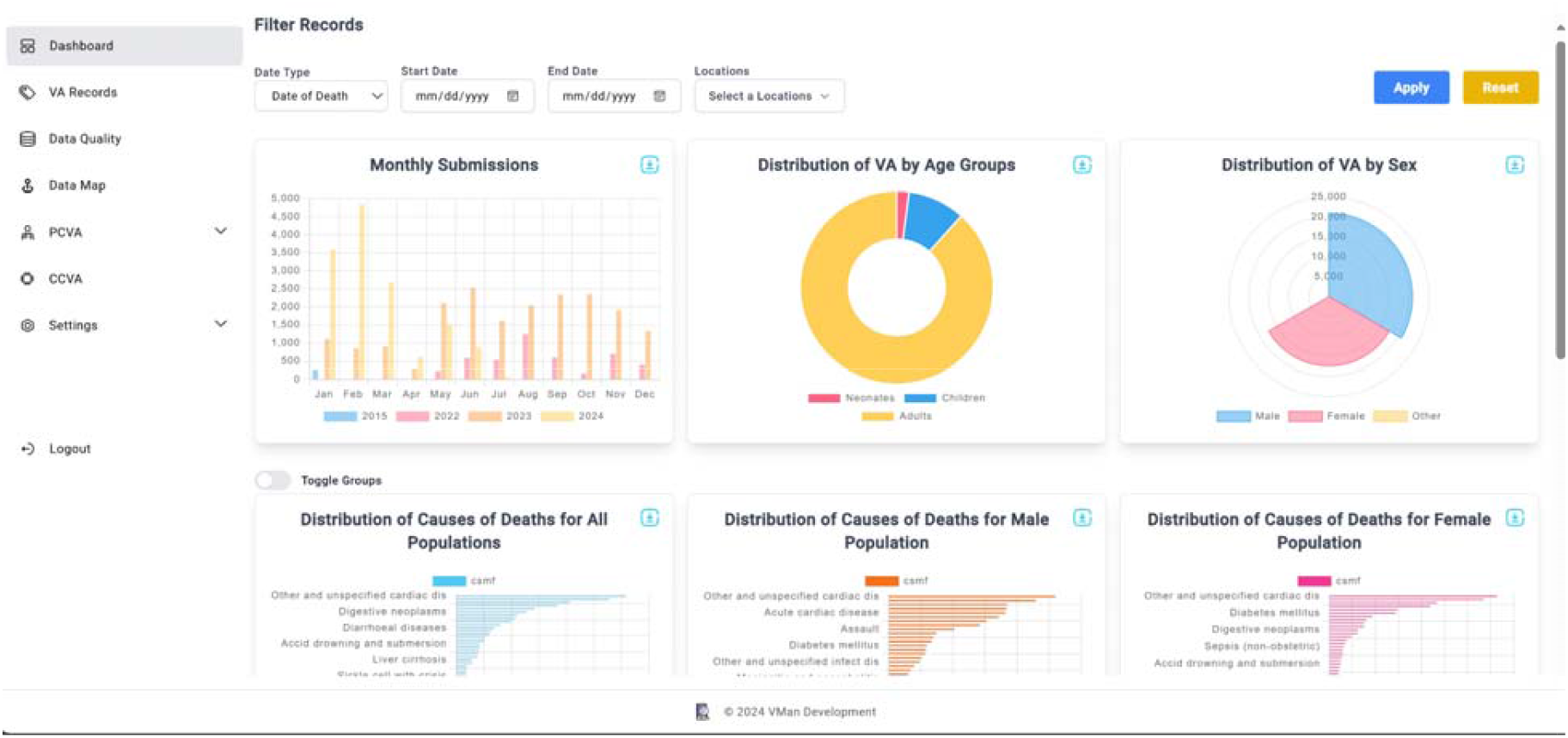
Dashboard module

### VA Records Module (Figure 3)

The VA Records Module contains a list of all VA records available in the system. This module allows the viewer to display individual contents of the VA document by clicking the eye-icon located on the right-hand side of the page. This functionality pulls all nodes including meta-data of the VA document, and displays the content in a pop-up window with vertical structure (Figure 3), allowing scrolling up and down for the entire record viewing capabilities. The right-hand side of the popped-up window contains a summary view of the VA. The selection of which items to display in the summary window is configured in the settings page.

**Figure 3:**
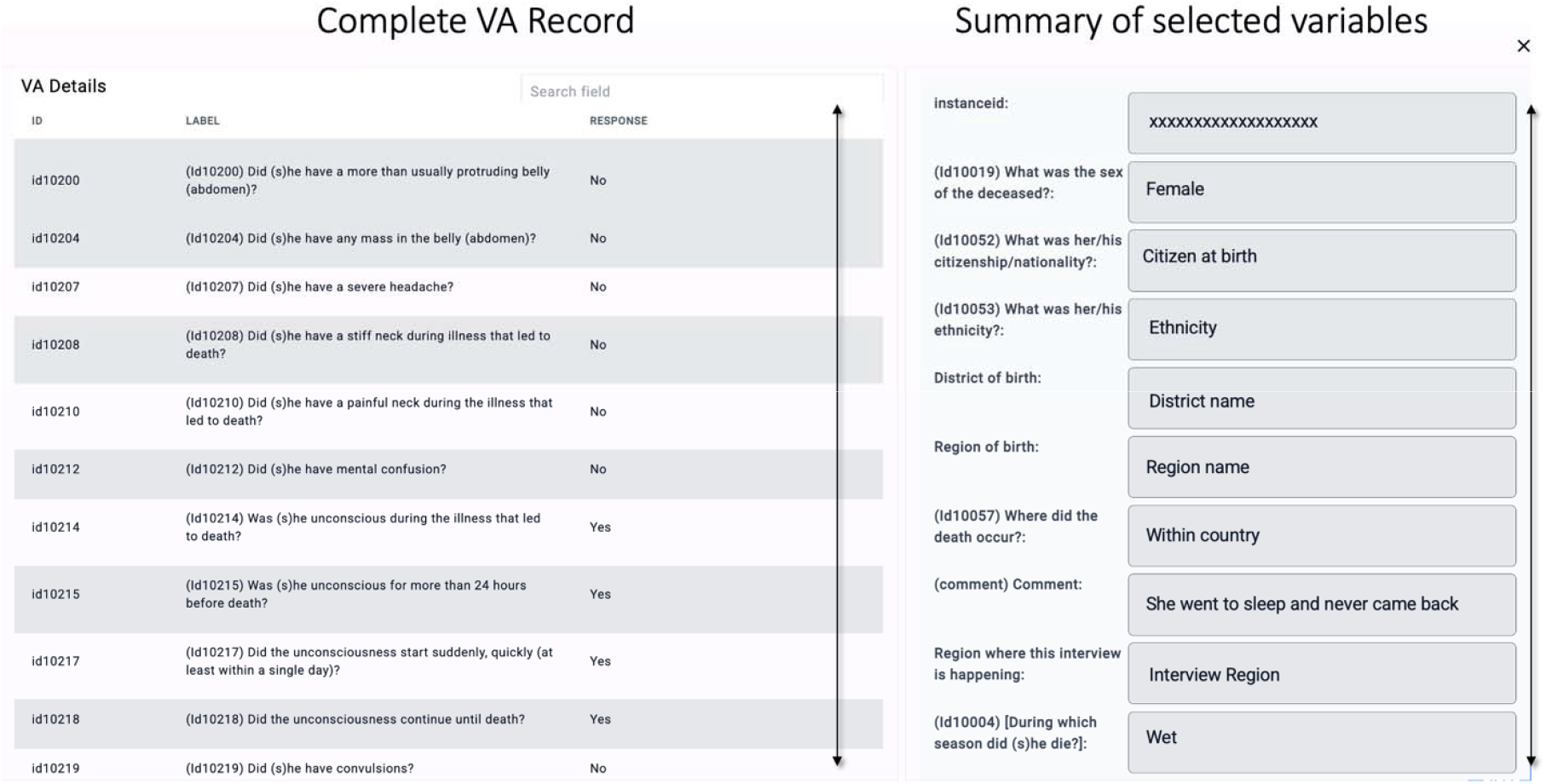
VA Records (Single record view) * The ID and LABEL fields are mapped using the ODK configuration file, while the RESPONSE is retrieved from the corresponding VA record in the database. Users can scroll vertically for additional visibility

### Global filter functionality

The global filter functionality is available on top of each of the following modules: Dashboard, VA records, and Data Map. The global filter gives the capabilities to folder records based on date (submission, interview or date of death) as well as interview location. When the global filter is applied on a given page, the content of the page will change to reflect choices which have been applied in the filter options.

### COD Determination

The VA records contain interview content which is structured as defined in the interviewing instrument. This content often contains history, signs and symptoms of the deceased in the period leading to death. It does not have information on the probable cause of death (COD). In order to establish the COD, the VA records need to be processed in either physician coding methods commonly referred as Physician Coded Verbal Autopsy (PCVA) or computer automated methods referred as Computer Coded Verbal Autopsy (CCVA). The VMan tool has both options, allowing easy processing of the COD data.

### Physician-Coded Verbal Autopsy (PCVA) Module

The Physician-Coded Verbal Autopsy (PCVA) module facilitates the assignment of cause of death (COD) by physician coders. When a user is assigned the role of a coder, they can be allocated Verbal Autopsy (VA) records for review and COD assignment. Upon logging into the system, an assigned physician coder can access VA records under the “Assigned VA” section. Selecting a record opens the coding interface (Figure 4), which consists of the VA document on the left-hand side and the coding sheet on the right-hand side. The coding sheet is adapted from the World Health Organization (WHO) 2016 Medical Certification of Cause of Death (MCCD) form (11). To streamline the process, the system automatically populates key demographic details—such as unique ID, age, and gender—into the coding sheet. Additionally, a notes section is available to enable coders to document observations for further reference.

**Figure 4:**
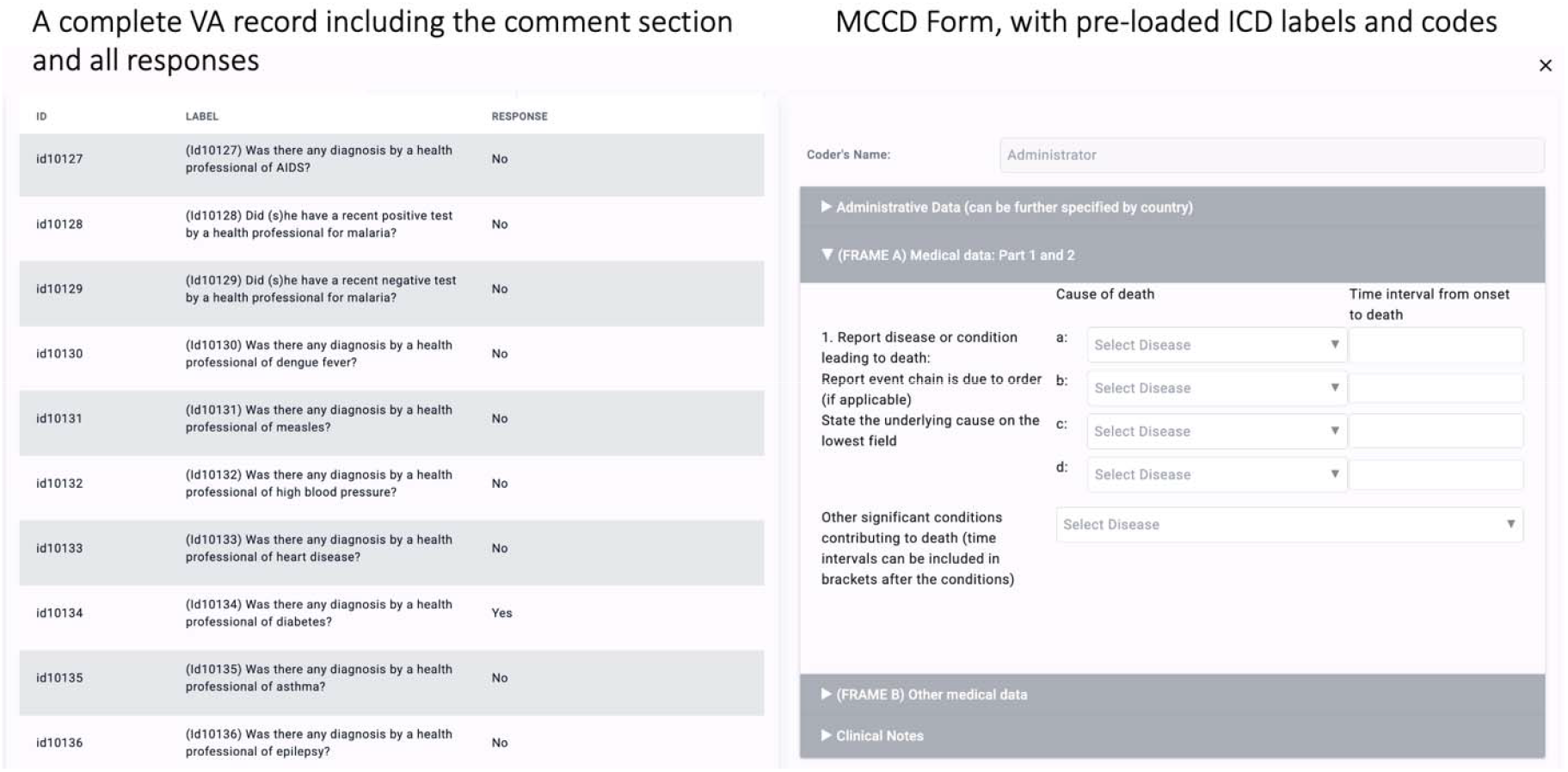
VMan coding window

**Figure 5:**
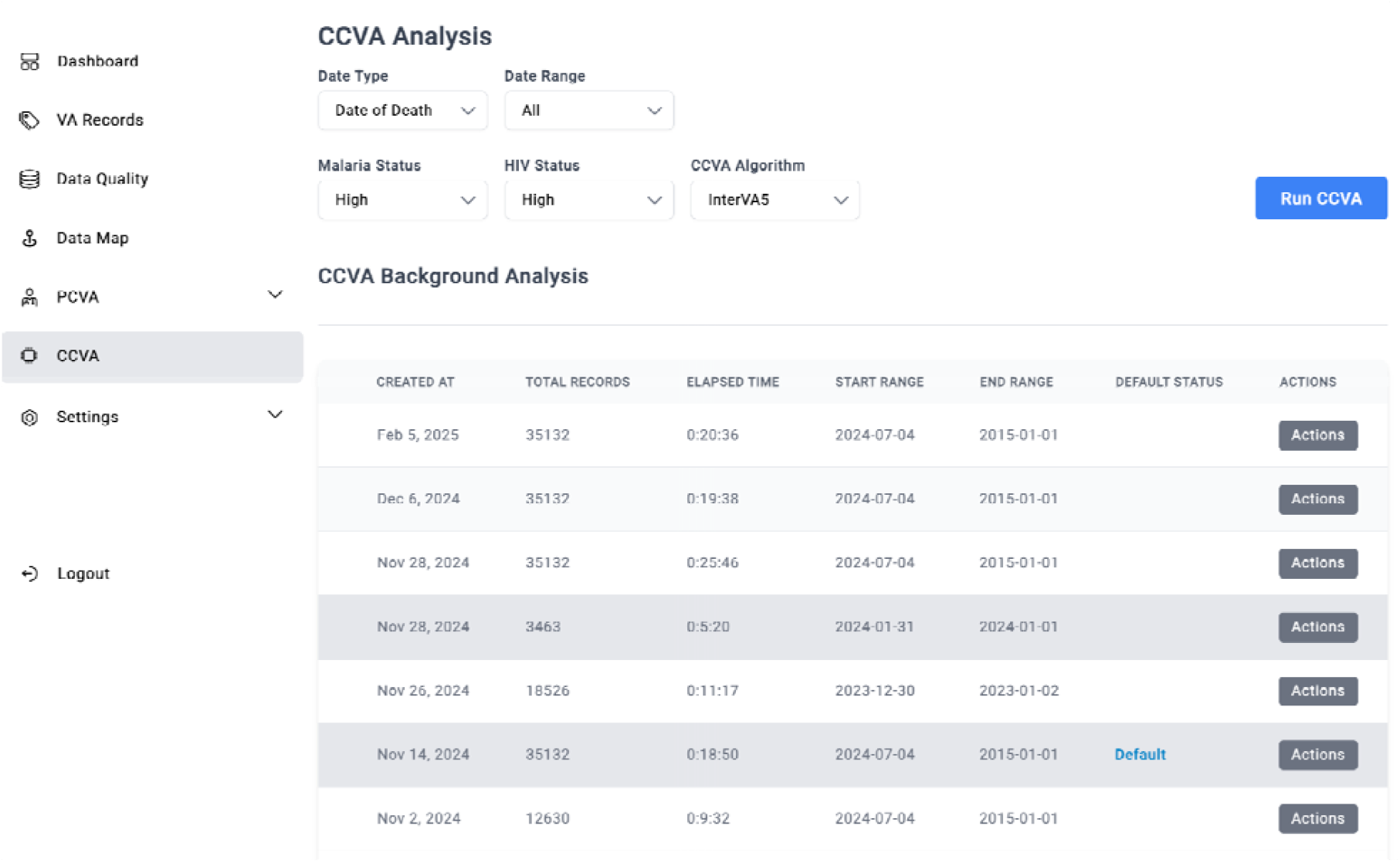
CCVA Module

### Integration of the International Classification of Diseases (ICD)

The system page allows configuration of ICD-10 lookup values. Provision for ICD-11 integration is made for future releases of the software. System administrators can import ICD-11 codes or manually enter ICD records, categorizing them as Communicable Diseases, Non-Communicable Diseases, or Injuries. Once configured, these standardized COD categories become available within the coding interface, ensuring consistency in cause-of-death classification.

Coding Agreement and Concordance: VMan3 allows a single VA record to be assigned to multiple coders, with the number of assigned coders configurable in the system settings. Coding agreement is determined based on the consistency of COD assignments:

1. **Concordat agreement** occurs when two or more coders independently assign the same underlying COD.
2. **Discordant agreement** arises when there is a discrepancy between COD assignments by different coders. The system automatically categorizes VA records into concordant and discordant groups for further review.

### Resolution of Discordant COD Assignments

For discordant cases, the system provides a Discordant Agreement Resolution Module. This module includes an integrated chat window, enabling coders to communicate and discuss discrepancies in COD assignment. Coders can exchange messages and review clinical evidence to reach a consensus. Once agreement is achieved, the record is reassigned to the concordant category, ensuring accuracy and consistency in COD classification.

### Computer Coded Verbal Autopsy (CCVA)

The Computer-Coded Verbal Autopsy (CCVA) module contains functionality for deriving COD to VAs using computer automated methods. These methods are a set of predefined libraries which can be imported into VMan and provide desired functionality. VMan is deployed with OpenVA InterVA5 library (12,13). During the CCVA processing, users can apply filters to select records based on specific date ranges or geographic locations. Users can generate multiple instances of CCVA-processed data, and select the most relevant dataset for displaying on the dashboard. While running the algorithms, apart from COD data, the CCVA module also produces data related errors which are found during data quality checks. These errors can be viewed in the system under the Data quality module. The data quality module allows users to browse each error and allows the opportunity to clean the record associated with the error. Cleaned records can be re-processed for COD determination.

### Data Sharing and Integration

VMan is built using FastAPI framework [ref], a modern, high-performance web-based framework for building Application Programming Interfaces (APIs) solutions. This approach facilitates a mechanism for secure sharing of VA data from VMan to other systems in the domain, including DHIS2 and civil registration systems. To ensure data security and compliance, access requests undergo secure authentication and validation processes.

### Test & Validation

The VMan software has been stress-tested with 38,000 VA records. The test configuration consisted of an instance of ODK Central (Version 1.4.2) with 38,000 records and an empty VMan instance. Both instances were deployed on Digital Ocean virtual machines with 16GB Ram, 4GB Intel vCPU and Ubuntu 23.10 x64 Operating System. Data synchronization took approximately 20 minutes. All records were synched successfully. The additional details and documentation of the VMan tool can be viewed at documentation page (https://www.vman3.vatools.net)

## Discussion

VA data is collected using ODK Collect, a mobile-based digital tool for data collection which is deployed with the ODK Systems. When submitted to the server, VA data is stored in the ODK database, and can be accessed digitally with the appropriate username and password. The latest release of the ODK (ODK Central) has a configurable API which allows easy pulling and pushing of the VA data. VA data is structured following the guidance of a standard WHO VA Instrument with allowable minor modifications in the demographics section to capture the setup of the geographic locations of the implementing country. This often causes minor differences in the data structure of VA data from different use-cases and VMan tries to resolve this problem by offering a harmonized universal data storage structure.

The tool is being designed for various personas including but not limited to project manager, data managers and statisticians, field supervisors, system administrators and physicians for conducting PCVA. In other setups epidemiologists can also utilize the algorithm output results to conduct further analysis and plausibility checks of the causes of death in relation to demographics in the VA data and for detection of PHEs. Furthermore, the system also allows export of data and graphs for use in further analysis. The PCVA section provides an electronic version of the standard MCCD form for use to capture physician entries when coding cases, which includes a dropdown list of the ICD 10 mortality Codes to facilitate accurate and quick coding. The ICD 10 mortality cause list is flexible and codes can be added or removed based on in-country recommendations.

Inorder to simplify the process of running cause of death algorithms, the CCVA module provides the user with variety of options to choose the algorithm to use, and the parameters required for the algorithm. The results can thereafter be exported in csv format for further analysis. By standardizing VA data management and automation, VMan will significantly contribute to strengthening mortality surveillance systems. It aligns with global efforts to improve mortality statistics and supports data-driven health policies, planning and PHE detection in LMICs.

## Challenges & Mitigation

The six-month funding period constrained the development process, providing a limited timeframe for implementation. Additionally, the funding did not support broader stakeholder engagement across multiple countries, restricting opportunities for co-creation and local ownership. To mitigate these challenges, virtual meetings and email communications were leveraged to facilitate collaboration and gather input.

## Conclusion

The development of VMan3 marks a significant advancement in verbal autopsy (VA) data processing and cause-of-death (COD) classification. Built on global reference architecture and WHO Smart Guidelines, VMan3 ensures standardized data collection, improved data quality, and seamless integration with national health information systems. Its adaptable architecture and modular design provide a robust foundation for addressing current challenges while supporting future innovations in VA implementation, including use of artificial intelligence (AI) to determine causes of death from community deaths. Countries and health projects are encouraged to adopt VMan3 to enhance VA data management, ultimately strengthening public health surveillance, policy decisions, and resource allocation.

## Acknowledgement

The authors gratefully acknowledge the support and collaboration of the Ministries of Health in Tanzania and Zambia, particularly for their active participation in technical meetings and contribution during the requirement-gathering sessions. We also extend our sincere appreciation to the AFENET team for the valuable assistance throughout project implementation and support during the preparation of this manuscript

## Declaration

All authors declare no conflict of interest.

## Ethics Statement

The test dataset was simulated using Tanzania’s CRVS-VA data collection tools. As this study focused exclusively on software development and testing without direct human subject involvement, Institutional Review Board (IRB) approval was not required. Location parameters and sample responses were derived from hashed, retrospective CRVS program data, ensuring no personally identifiable information was included in the simulated dataset.

## Author contribution

IL led the conceptualization and preparation of the initial manuscript draft. CO, GM, SM, and SM provided critical review and contributed contextual insights relevant to the local implementation. CO also curated and added references. JM and CM were responsible for developing the software’s source code and contributed to the interpretation of its functionalities. SI revised the visual content and enhanced the documentation with additional references. CO further revised the background section and expanded the reference base. RM and GK reviewed the final document and provided overall feedback to finalize the manuscript.

## Data availability

The test dataset for this study was generated using the WHO 2016 Verbal Autopsy (VA) Instrument, which is publicly available on the World Health Organization (WHO) website (5). While other versions of the WHO VA instrument may be compatible, they have not been validated with this software.

